# Cardiovascular Risk Models in Chronic Kidney Disease: A Systematic Review and External Validation of Models

**DOI:** 10.1101/2022.01.22.22268958

**Authors:** Rupert W Major, Robert Grant, David Shepherd, James F Medcalf, Jorge Jesus-Silva, Laura J Gray, Nigel J Brunskill

**Author notes:** Corresponding Author: Dr Rupert W Major, Department of Health Sciences, George Davies Centre, 15 Lancaster Road, University of Leicester, Leicester, UK.

## Abstract

**Background and Objectives:** Risk factors for cardiovascular disease in chronic kidney disease differ to the general population due to the increased role of inflammation, calcification and arteriosclerosis. Over 350 cardiovascular risk models (“models”) exist in the general population but most have not undergone testing (“external validation”) in other populations, such as cohorts with chronic kidney disease. We aimed to update a previous systematic review of models in chronic kidney disease and then perform external validation of these and general population models in a chronic kidney disease cohort.

**Design, Setting, Participants and Measurements:** We searched Medline up to 26^th^ August 2020 for models in chronic kidney disease. We performed external validation of models using a primary care chronic kidney disease cohort of 17,248 individuals with 2,072 (12.0%) cardiovascular events, 5,108 (29.6%) deaths and a median follow-up of 5.0 years. Model discrimination and calibration was assessed and where appropriate models were re-calibrated. Multiple imputation was used to account for missing data.

**Results:** Seven chronic kidney disease specific models were identified. These models and three general population models underwent external validation. All models had worse discrimination of events than in their original cohorts, particularly in the chronic kidney disease specific models. General population models were miscalibrated and overpredicted risk. The major contributor to this was the high competing risk of death from non-cardiovascular causes.

**Conclusions:** Existing chronic kidney disease cardiovascular risk models performed poorly in external validation. General population cardiovascular disease risk models should be interpreted with caution in individuals with chronic kidney disease as they may overestimate risk due to the competing risk of death. Further development of models to include simultaneous risk prediction for cardiovascular and non-cardiovascular disease risk is required.

## Introduction

Chronic kidney disease (CKD) is independently associated with increased risk of cardiovascular (CV) disease (1). Risk factors for CV disease in those with CKD may differ from ‘traditional’ CV risk factors found in a general population, due to the role of inflammation, calcification and arteriosclerosis in CKD, particularly with more advanced disease (2).

Therefore, specific CV risk prediction models for use in those with CKD may be warranted. In 2013 Tangri *et al* identified five CV risk models developed in CKD cohorts, but noted their limitations, including their lack of external validation in independent cohorts (3). The models were also developed in secondary care cohorts despite most individuals with CKD being managed in primary care. Most primary care CKD is at low risk of development of endstage kidney disease and CV risk management and prediction is the more pertinent issue.

There are more than 350 general population CV risk models, most of which are not used or integrated into routine patient care (4). Those that are used focus on prediction in primary prevention populations and most did not consider CKD in their development, despite substantial evidence for its association with CV disease. From those recommended at a national or international level only QRisk3 includes the presence of CKD stage 3, 4 or 5 as a binary predictive variable based on primary care coded CKD (5). However, up to a third of coded CKD may be incorrect and therefore brings into question its validity in people with CKD (6). Given that the inclusion of estimated glomerular filtration rate (eGFR) and quantification of proteinuria through albumin to creatinine ratio (ACR) in CV risk prediction has also been demonstrated (7), specific model assessment in CKD is justified.

External validation improves the generalisability of risk models and may indicate their suitability for implementation into clinical practice, although this is rarely performed (8). Effectively implemented risk models can aid patient choice in relation to treatment and prognostication, as well as supporting more effective clinical research, such as clinical trials, and health care system management (8).

The first aim of the current manuscript was to identify additional CV risk models in CKD published since Tangri *et al*’s review. Secondly, we aimed to perform an independent, external validation of CKD specific and general population CV risk models identified in a primary care CKD cohort.

## Materials and Methods

### Literature Review – Identification of Prediction Models

In order to update the previous systematic review of CV risk prediction models in CKD, Ovid MEDLINE was searched from 2012 to 26^th^ August 2020. The Search strategy was based on Tangri *et al*’s literature review terms (3). The title and abstracts of all studies identified by the literature search were assessed. The full text of any abstract meeting the inclusion criteria was then reviewed in full. All stages of the literature review, data extraction and bias assessment were performed independently by two reviewers. All identified models had core characteristics extracted including definitions and number of CV events and the hazard ratios and/or beta-coefficients for variables included in the model. Specifically for the risk prediction models baseline hazard, discrimination and calibration metrics were also extracted. General population models were selected on the basis of their recommendation in national and international CV guidelines for North America and Europe.

### Cohort Description for External Validation

‘The Leicester City and County Chronic Kidney Disease’ (LCC) (ClinicalTrials.gov Identifier: NCT03135002) cohort was used for external validation of eligible risk models (9). Ethical approval was received from the national agency for approval of healthcare research in the UK (‘Health Research Authority’). Adult individuals were included in the cohort if they had two or more CKD-EPI eGFRs <60 ml/min/1.73m^2^ >90 days apart prior to the start of the follow-up period of 1st November 2011. Co-morbidities were defined using the same criteria as the CKD Prognosis Consortium (7). CV events were identified from primary and secondary care records up until 1^st^ November 2016. Individuals with a kidney transplant or on dialysis at the baseline date were excluded.

### Statistical Methods

All models were assessed for potential external validation. Models were eligible if their outcome included fatal and non-fatal CV events. The full detail of risk models, including their baseline risk, are not always presented therefore the “level” of published information was assessed (10). Level one data refers to the publication of regression coefficients only. Level two includes level one plus risk groups and associated Kaplan-Meier plots. Level three includes the previous levels of data plus the baseline hazard or survival function. The different levels of data allow different model metrics to be calculated from the validation dataset. Only level three data allows for assessment of calibration of the model.

Multiple imputation of missing values of variables in models was performed. Imputed values were then used to calculate predicted risk before assessment of models was performed. One imputation cycle was performed for every percent of incomplete values for a variable, which is a conservative prediction for the fractional of missing information (11). Sensitivity analysis using complete cases only was also performed.

Harrell’s C-statistic was used to assess general discrimination for all models. Where Level two data were available, Kaplan-Meier curves for risk groups were plotted and hazard ratios across risk groups calculated. Calibration was assessed where level three data were presented using calibration plots. Risk groups were based on deciles of calculated risk in the original model. All risk calculations were based on the full baseline risk reported for a model, with adjustment of the baseline risk to the five year timeframe of the LCC cohort. Baseline risk adjustment was performed using a proportional method with sensitivity analysis performed using an exponential method for adjustment. All assessments were made using the whole LCC cohort (‘whole cohort’) and the subgroup of the LCC cohort matched to the characteristics and outcome of the cohort that the CV risk model being assessed was developed for (‘cohort specific’). For example, if a risk model had been developed for primary prevention of CV disease then only individuals without a history of CV disease were included in the cohort specific model assessment. All results are reported in line with the “Preferred Reporting Items for Systematic Reviews and Meta-Analyses” (PRISMA) and the “Transparent reporting of a multivariable prediction model for individual prognosis or diagnosis” (TRIPOD) statements (Supplementary Items 1 and 2) (12). In relation to patient and public involvement in this research, the grant to perform the study underwent review by patient representatives before submission to the funding body and by a separate group of patient representatives during their review and approval of the grant. All statistical analysis was performed using Stata 16.0 (StataCorp LLC, TX, USA), with p<0.05 considered statistically significant.

## Results

### Updated Literature Review

#### CKD Models

Supplementary Figure 1 shows the screening process, including the number of publications and risk models identified, and reasons for any exclusion. 4,493 abstracts were identified on Medline and 44 full-text articles were reviewed. The full search strategy is shown in Supplementary Item 3. Seven studies were identified, presenting eight new models. Therefore, including the models identified by Tangri *et al*, 16 models were assessed for potential external validation, of which seven were suitable for external validation (13,14,15). Three models could not be externally validated as they presented CV mortality data only and six contained continuous variables, such as research biomarkers, that were not available in the external validation cohort’s routinely collected data.

Two of the cohorts excluded individual with existing CV disease at baseline. The models’ characteristics are shown in Table 1. Similarly to Tangri *et al*’s earlier findings, presentation of models was suboptimal (3). None of the new models presented all core model metrics of discrimination, calibration, model fit, reclassification and external validation. Level three data were available for four out of the five models. Three of these models used a baseline hazard function from another study, the Framingham Heart Study, and did not specify a re-calibrated baseline function for their own model. Variables included in the models are shown in Table 2. All models included age, gender, smoking, diabetes mellitus and hypertension. One model included CV disease and the other four excluded individuals at baseline with these conditions. eGFR was included in two models, one of which also included a measurement of proteinuria. No other models included proteinuria as a risk factor.

**Table 1:**
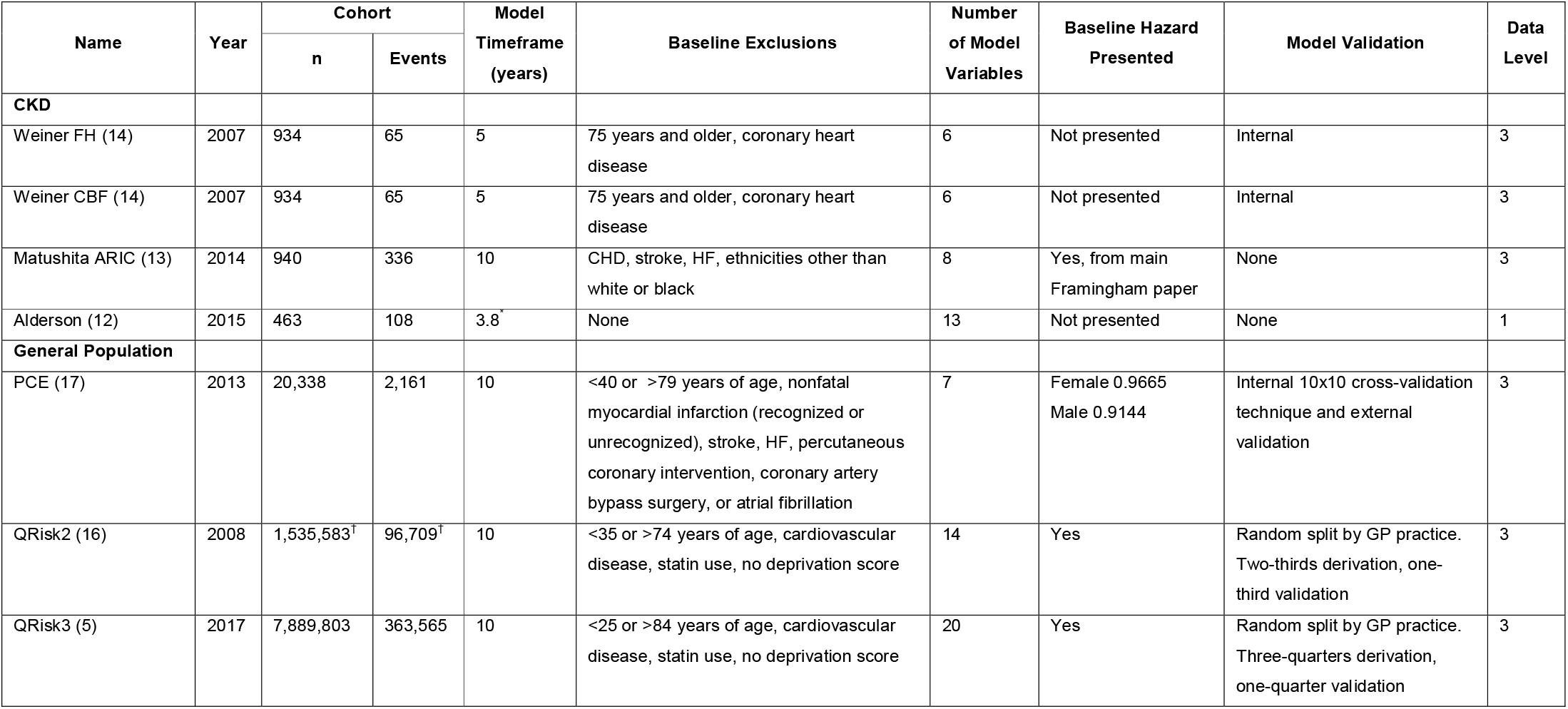
Summary Characteristics of Risk Models for External Validation. Based on findings of Tangri *et al* and updated literature review. * - median follow-up, † - refers to derivation cohort.

**Table 2:** Variables included in identified models. N/A – no individuals with CVD in cohorts. * - separate models with same variables presented for male and female cohorts. TC – total cholesterol, HDL – high density lipoprotein, BNP – brain natriuretic peptide, PTH – parathyroid hormone

| Model | Age | Gender | Smoking | CVD | DM | HTN | eGFR | Proteinuria | TC | HDL | Other |
| --- | --- | --- | --- | --- | --- | --- | --- | --- | --- | --- | --- |
| Weiner – FH (14) | • | •* | • | N/A | • | • |  |  |  |  |  |
| Weiner – CBF (14) | • | •* | • | N/A | • | • |  |  |  |  |  |
| Matushita – ARIC (13) | • | • | • | N/A | • | • |  |  | • | • | BNP |
| Alderson (12) | • | • | • | • | • | • | • |  |  |  | Serum phosphate, calcium, albumin, haemoglobin, PTH |
| PCE (17) | • | •* | • | N/A | • | • |  |  | • | • | Interaction terms include – age*TC, age*HDL, age*SBP, age*smoking |
| QRisk2 (16) | • | •* | • | N/A | • | • |  |  | • | • | BMI, family history of CHD, deprivation, treated hypertension, rheumatoid arthritis, CKD, AF |
| QRisk3 (5) | • | •* | • | N/A | • | • |  |  | • | • | BMI, family history of CHD, deprivation, treated hypertension, rheumatoid arthritis, CKD, AF, SBP variability, migraine, corticosteroid use, systemic lupus erythematosus, 2<sup>nd</sup> generation “atypical” antipsychotic use, severe mental illness, erectile dysfunction |

#### General Populations Models

Four main models were identified: The American Heart Association Pooled Cohort Equation (PCE), The European Society of Cardiology (SCORE) and National Institute of Health and Clinical Excellence (England and Wales) (QRisk2 and QRisk3) (5,16,17,18). All general population models produced specific models for males and females and these were therefore assessed separately. PCE also produced separate models for non-Hispanic black populations, however, because the cohort used for assessment only included 14 events from the 123 black individuals included, these models were not assessed. SCORE only assessed CV mortality outcomes and so was not externally validated.

#### Brief Cohort Description

One hundred and three primary care practices were invited to participate in the LCC cohort, of whom 44 (42.7%) agreed. These practices represented a total adult population of 277,248 individuals. The cohort consisted of 17,248 (6.2%) individuals with two or more CKD-EPI eGFRs <60 ml/min/1.73m^2^ >90 days apart prior to the start of the follow-up period. 10353 (60.0%) individuals were female and mean age was 77.4 (SD 10.0) years. Median CKD-EPI EGFR was 52 (IQR 43 to 56) ml/min/1.73m^2^ and median ACR was 2.9 mg/mmol (IQR 0.8 to 7.3). 12365 (71.7%) of the cohort had CKD eGFR stage 3a and 5,846 (33.9%) had ACR stage A1. Table 3 describes the cohort in further detail and Supplementary Table 1 describes the cohort according to KDIGO CKD stage.

**Table 3 -.** Baseline characteristics of the LCC cohort. BP – blood pressure, HDL – high density lipoprotein cholesterol, LDL – low density lipoprotein cholesterol. Units – eGFR ml/min/1.73m^2^, ACR and lipid mg/mmol, BP mmHg.

| Variable | Whole Cohort |  | Missing Data |  |
| --- | --- | --- | --- | --- |
|  | Mean/Median/n | SD/IQR/% | n | % |
| Age in years | 77.4 | 10.0 | 0 | 0.0 |
| Female | 10,353 | 60.0% | 0 | 0.0 |
| White Ethnicity | 12,560 | 72.8% | 4,687 | 13.7 |
| Former Smoker | 1,421 | 8.2 | - | - |
| Ex-smoker | 6,256 | 36.3 | - | - |
| Mean ACR | 10.6 | 35.4 | 5,614 | 32.6 |
| Median ACR | 2.9 | 0.8 to 7.3 | 5,614 | 32.6 |
| Mean CKD-EPI eGFR | 48.5 | 9.9 | 0 | 0.0 |
| Median CKD-EPI eGFR | 52 | 43 to 56 | 0 | 0.0 |
| Diabetes Mellitus | 5,592 | 32.4% | - | - |
| Hypertension | 15,674 | 90.9% | - | - |
| Systolic BP | 134.8 | 16.4 | 195 | 1.1 |
| Diastolic BP | 74.0 | 9.9 | 104 | 0.6 |
| Ischaemic Heart Disease | 4,384 | 25.4% | - | - |
| Heart Failure | 1,690 | 9.8% | - | - |
| Cerebrovascular Disease | 2,127 | 12.3% | - | - |
| HDL | 1.4 | 0.4 | 1,214 | 7.0 |
| LDL | 2.4 | 0.9 | 2,009 | 11.6 |
| Total Cholesterol | 4.6 | 1.1 | 763 | 4.4 |
| Mean Deprivation Decile | 5.7 | 3.0 | 362 | 2.1 |
| Median Deprivation Decile | 6 | 3 to 8 | 362 | 2.1 |

2072 (12.0%) CV events occurred over a median follow-up of 5.0 (IQR 3.3 to 5.0) years. 5108 individuals (29.6%) died during follow-up and 155 (0.9%) required either dialysis or a kidney transplant. 11,410 (66.2%) had no previous coded history of ischaemic heart disease or cerebrovascular disease at the study’s baseline. 1,035 (9.1%) of this group has a CV event during follow-up.

### Model Performance

#### Discrimination

Overall discrimination for the identified CKD and general population models is shown in Table 4 for the summary imputed results and for complete case analysis in Supplementary Table 2. In general, all models had worse discrimination in the current cohort than in their original development models. The Matsushita Framingham based model had the best discrimination of the CKD based models. However, overall CKD based models performed worse than general population models both in the current whole cohort and cohort specific group. Results were similar for complete case analysis.

**Table 4:** Summary C-statistics for CKD and General Population CV prediction models. C-statistics and 95% CI presented are the mean of c-statistic results for each imputed cycle. C – Harrell’s concordance statistic, CI – confidence interval.

| Model | Original Performance |  | Cohort Specific |  | Whole Cohort |  |
| --- | --- | --- | --- | --- | --- | --- |
|  | C | 95% CI | C | 95% CI | C | 95% CI |
| **CKD** |  |  |  |  |  |  |
| Alderson (12) | 0.873 | 0.825 to 0.921 | n/a | n/a | 0.652 | 0.641 to 0.664 |
| Weiner Male Framingham (14) | 0.62 | not reported | 0.577 | 0.556 to 0.599 | 0.569 | 0.554 to 0.585 |
| Weiner Male Best Cox (14) | 0.72 | not reported | 0.560 | 0.533 to 0.586 | 0.564 | 0.546 to 0.583 |
| Weiner Female Framingham (14) | 0.77 | not reported | 0.549 | 0.531 to 0.568 | 0.553 | 0.537 to 0.569 |
| Weiner Female Best Cox (14) | 0.82 | not reported | 0.542 | 0.523 to 0.562 | 0.542 | 0.525 to 0.559 |
| Matsushita Male Framingham (13) | 0.679 | not reported | 0.645 | 0.613 to 0.677 | 0.591 | 0.572 to 0.610 |
| Matsushita Female Framingham (13) | 0.679 | not reported | 0.610 | 0.586 to 0.633 | 0.601 | 0.585 to 0.617 |
| **General Population** |  |  |  |  |  |  |
| QRisk2 Male (16) | 0.792 | 0.789 to 0.794 | 0.634 | 0.579 to 0.688 | 0.625 | 0.606 to 0.643 |
| QRisk2 Female (16) | 0.817 | 0.814 to 0.820 | 0.670 | 0.633 to 0.707 | 0.656 | 0.641 to 0.671 |
| QRisk3 Male (5) | 0.858 | 0.857 to 0.860 | 0.628 | 0.573 to 0.683 | 0.618 | 0.599 to 0.637 |
| QRisk3 Female (5) | 0.880 | 0.879 to 0.882 | 0.684 | 0.647 to 0.720 | 0.666 | 0.651 to 0.681 |
| PCE Male (17) | 0.746 | not reported | 0.667 | 0.617 to 0.717 | 0.621 | 0.603 to 0.639 |
| PCE Female (17) | 0.806 | not reported | 0.645 | 0.607 to 0.684 | 0.665 | 0.650 to 0.680 |

In relation to the general population models, the QRisk3 model for the female population (‘QRisk3 Female’) had the best discrimination. For male population models, the PCE model had the highest c-statistic but it was not significantly different to the other male general population models. Therefore, the QRisk3 Female and Male models were selected for sensitivity analysis of their discrimination and for assessment of calibration.

Supplementary Table 3 shows the results of the sensitivity analysis for discrimination for QRisk3. Generally, model discrimination performed better in lower age groups, with the caveat of smaller sizes limiting their interpretation. Discrimination was similar for the separate outcomes of myocardial infarction only, stroke only, death only and the composite outcome of either a cardiovascular event or death. There were minor improvements in discrimination performance when the optimal beta coefficients for eGFR and ACR were added to the linear predictor of the QRisk models.

### Assessment of Calibration

Due to the relatively poor discrimination performance of CKD specific models compared to the general population models, calibration was only considered for the latter. The general population models, PCE Female and Male models, and QRisk3 Female and Male models, all overpredicted risk for the Whole Cohort analysis (Figures 1a,2a,3a,4a). Similar levels of overprediction occurred in the Cohort Specific analyses (Figures 1b,2b,3b,4b). Re-calibration of the baseline survival improved calibration for both the Whole Cohort analyses (Figures 1c,2c,3c,4c) and the Cohort Specific analyses (Figures 1d,2d,3d,4d). The new estimated baseline survival was slightly higher for the Cohort Specific analyses, for example in the QRisk3 Female analysis a 28.6% increase for the Whole Cohort and 44.6% increase for the Cohort Specific group.

**Figure 1:**
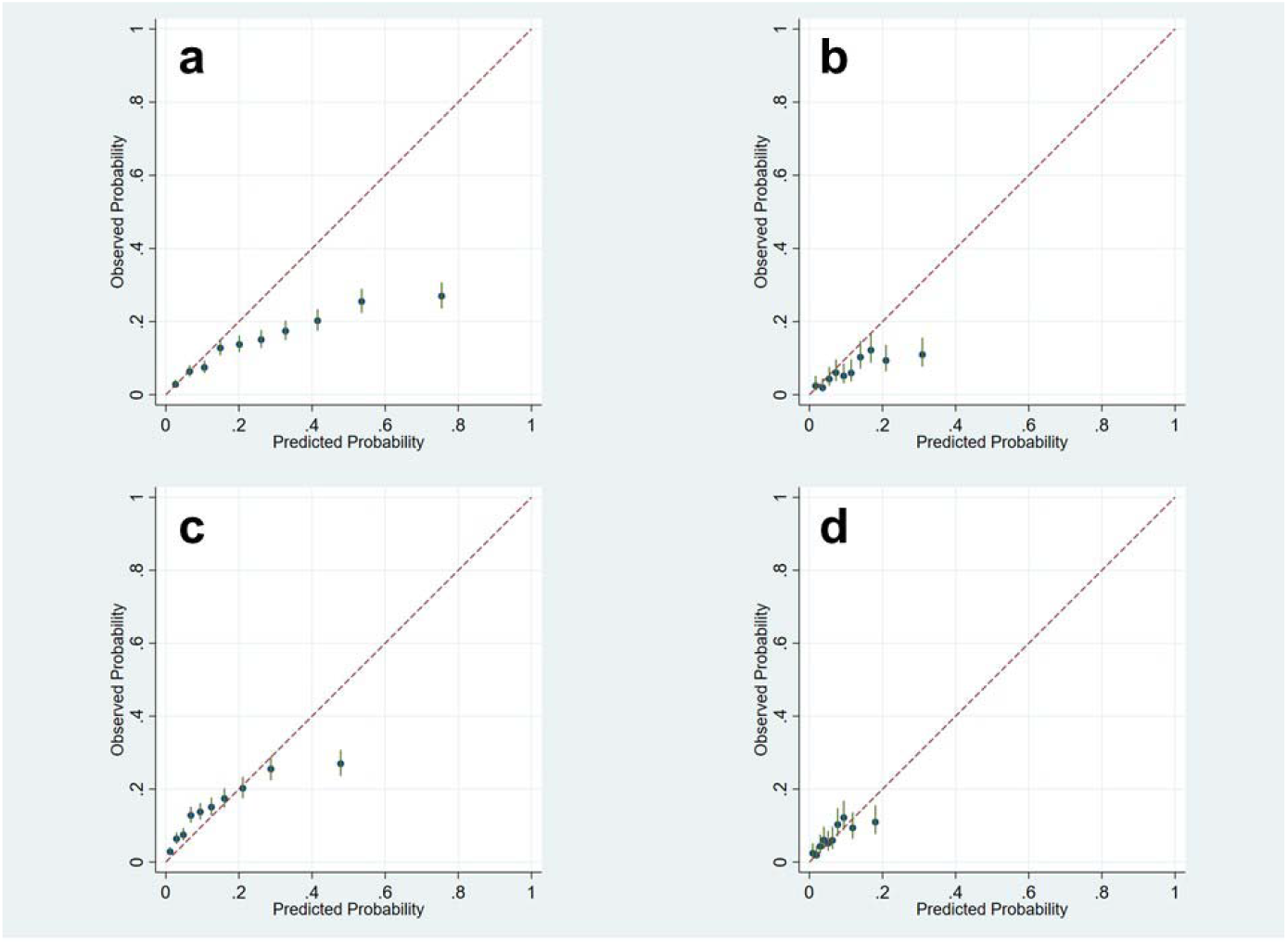
Calibration plots for **PCE Female** model for Whole Cohort (2a) and Cohort Specific (2b) groups. The lower half of the figure shows re-calibrated models for the Whole Cohort (2c) and Cohort Specific (2d) groups. Groups are split in to risk deciles.

**Figure 2:**
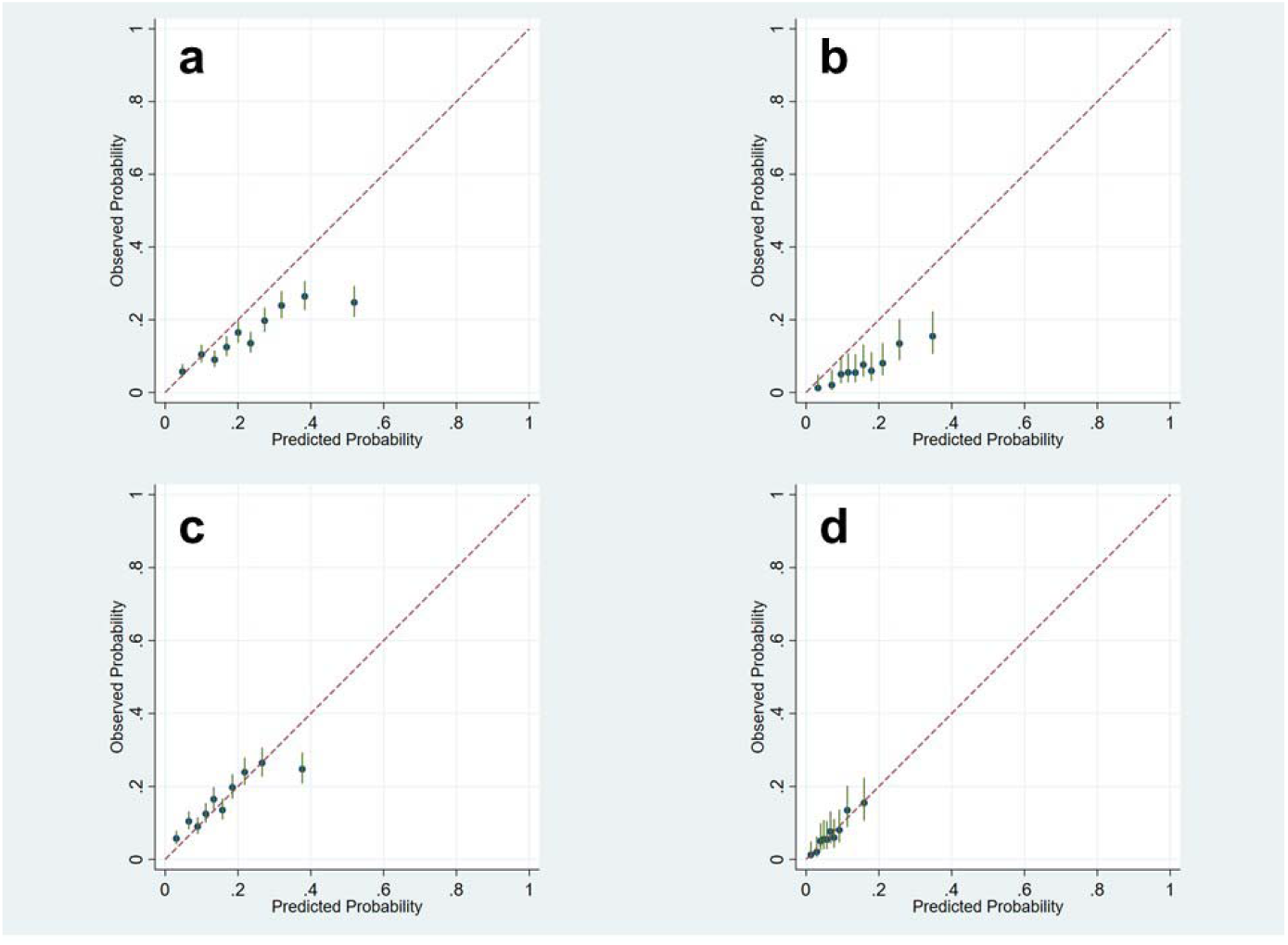
Calibration plots for **PCE Male** model for Whole Cohort (3a) and Cohort Specific (3b) groups. The lower half of the figure shows re-calibrated models for the Whole Cohort (3c) and Cohort Specific (3d) groups. Groups are split in to risk deciles.

**Figure 4:**
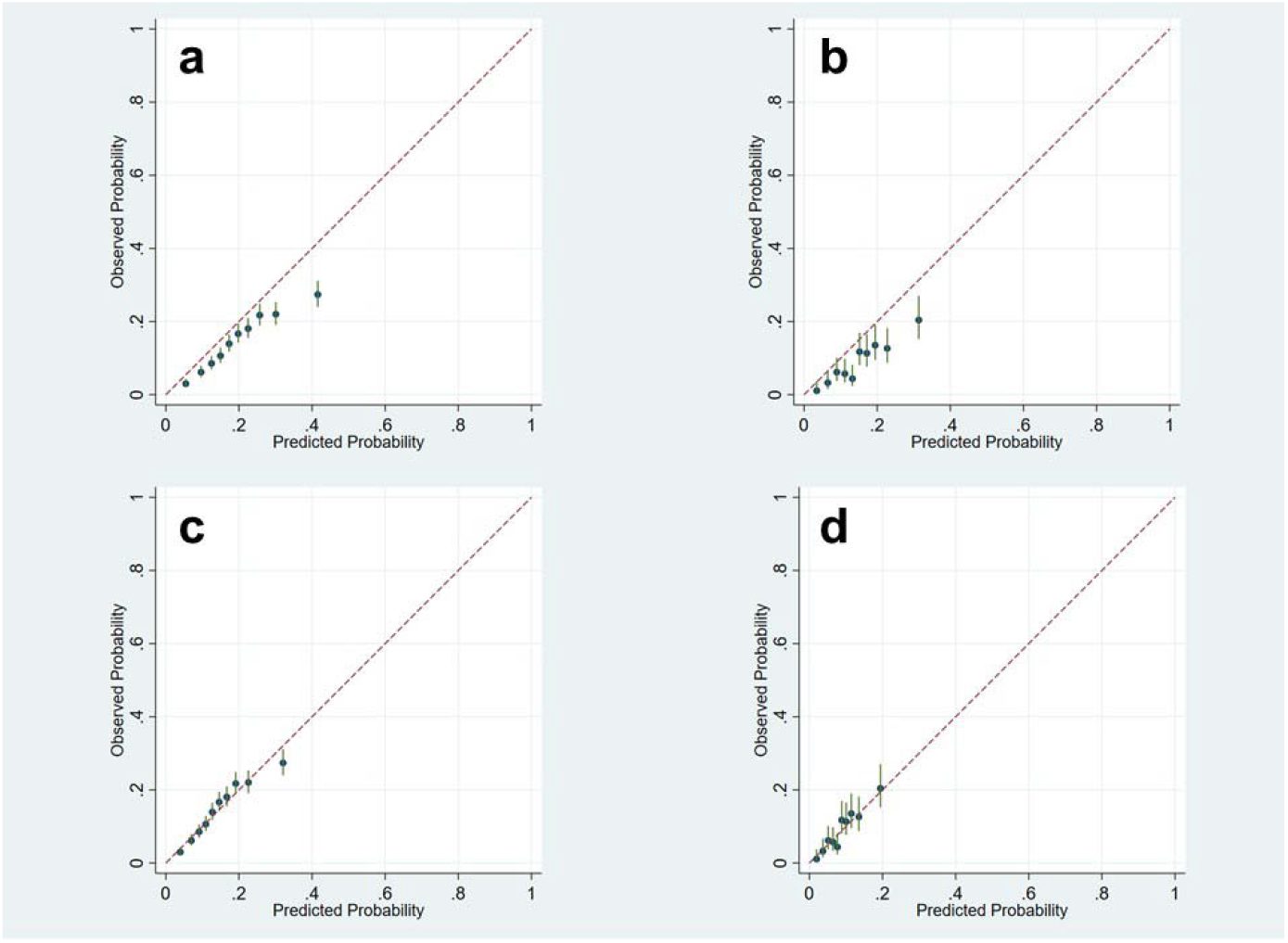
Calibration plots for **QRisk3 Female** model for Whole Cohort (4a) and Cohort Specific (4b) groups. The lower half of the figure shows re-calibrated models for the Whole Cohort (4c) and Cohort Specific (4d) groups. Groups are split in to risk deciles.

**Figure 5:**
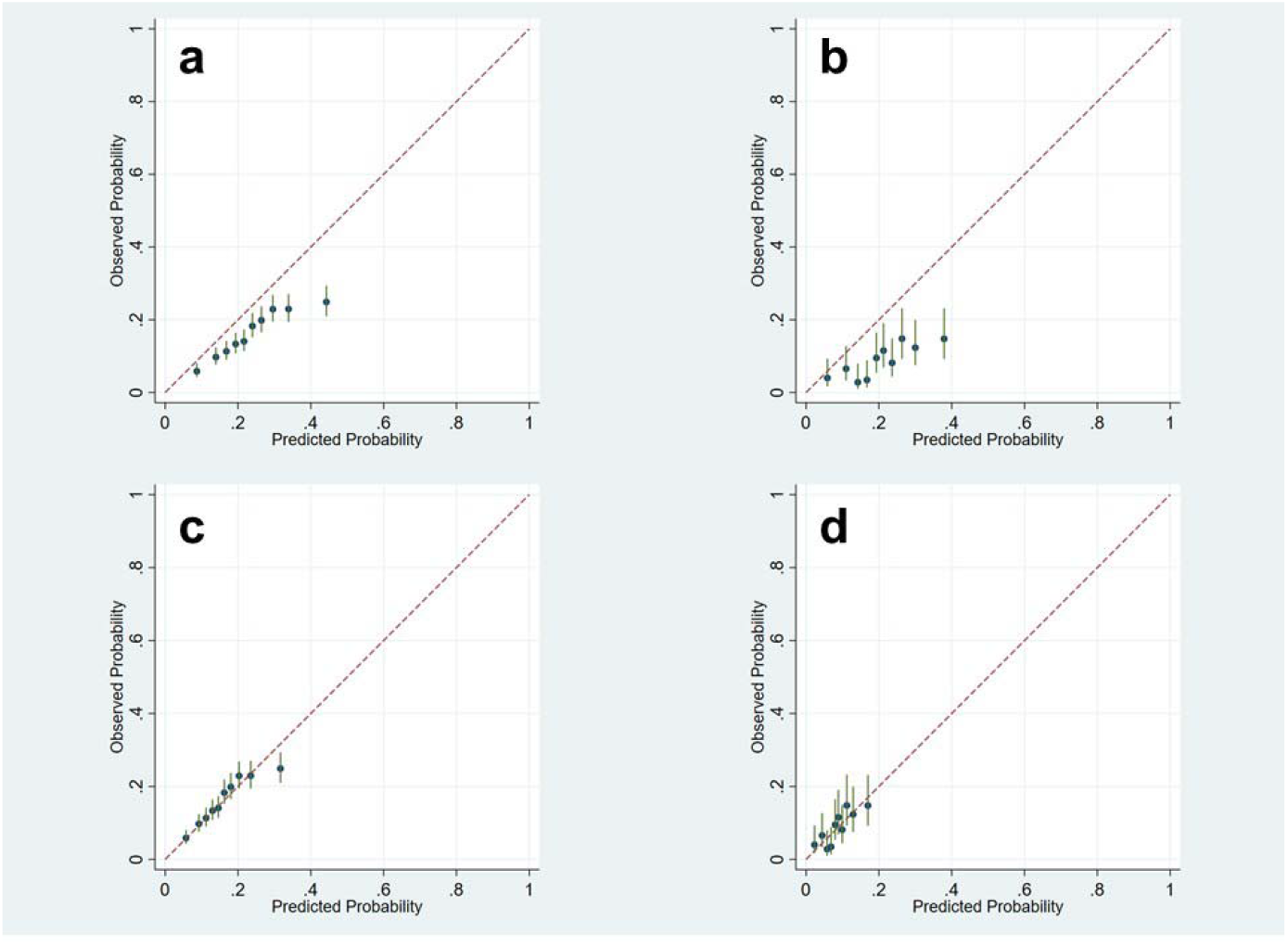
Calibration plots for **QRisk3 Male** model for Whole Cohort (5a) and Cohort Specific (5b) groups. The lower half of the figure shows re-calibrated models for the Whole Cohort (5c) and Cohort Specific (5d) groups. Groups are split in to risk deciles.

## Discussion

CKD is independently associated with increased risk of adverse outcomes, including CV events (1,19). There are over 350 CV risk prediction models and many have poor methodology and are probably not suitable for clinical use (4). Most have not been specifically externally validated in CKD populations. CV risk factors may be different in CKD compared to the general population both in terms of different magnitudes of effects of traditional risk factors and the role of non-traditional risk factors related to arteriosclerosis, cardiomyopathy and inflammation (2).

The current manuscript aimed to update a systematic review of cardiovascular risk prediction models in CKD and then externally validate them using a large CKD cohort. The update to the previous systematic review by Tangri *et al* (3) identified three additional new models from two cohorts. No publications of external validation of existing models was identified. These new models had similar limitation to those previously identified. In addition, suitable CV models used in the general population were also identified through review of international guidelines. The LCC cohort was then used to externally validate existing CV disease risk models.

Model discrimination was generally lower in all CKD models compared to general population models, with the best performing CKD model showing a C-statistic of 0.65. Worse discrimination in external validation is a common finding in external validation and does not necessarily invalidate the use of the risk prediction model in clinical care (20). For the general population models, QRisk models probably performed better in the female population and the PCE model better in males. Results were similar where the exclusion criteria of the development models were used to exclude individuals from the LCC cohort. Despite the smaller cohort for these specific analyses there continued to be an appropriate number of events to perform external validation (21). Predicted risk was probably higher, based on the mean linear predictor values, in the LCC cohort compared to the tested models. This is likely to reflect a more co-morbid population in the LCC cohort compared to general population groups.

As general population models showed equal or superior performance to CKD specific models, calibration was assessed in detail in the general population models only. These models generally overpredicted risk which was partially correctable by re-calibration. In sensitivity analysis, there were some minor improvement to model performance through the addition of eGFR and ACR which were of a similar magnitude to those previously described. (14).

This study has a number of strengths. Updating a previously used systematic review meant that all suitable models were considered in the external validation of models. The cohort used for external validation was a large CKD cohort with over 17,000 individuals and was specifically created for the purpose of assessment of risk prediction models. Over two thousands CV events occurred during five years of follow-up for the cohort which should be more than adequate to perform appropriate external validation (21). Identification of CV events was also robustly performed through data linkage between primary and secondary care. Due to the clinical nature of the data, multiple imputation was used to account for missing data to increase statistical power of the study. The point estimates for complete case analysis did not vary from the imputed analysis.

This manuscript has some limitations. Assessment of the QRisk3 models were limited by some categorical variables not being available in the LCC cohort. These were mostly low prevalence factors such as the presence of systemic lupus erythematous or “severe” mental illness.

The major finding of mis-calibration may be related to a number of cohort and methodological reasons. The baseline hazard was adjusted to reflect the five year timeframe of follow-up for the LCC cohort compared to the ten year prediction window for the general population time frame. Whilst this may possibly explain some of the mis-calibration, it is unlikely to explain all of this phenomenon. Missed identification of CV events may have occurred but would have been minimised through data linkage. Both of these potential contributors were investigated and not found to be a major cause of mis-calibration. Perhaps the major methodological factor relates to the limitation of the Cox proportional hazards models used to develop all of the identified models. One of its major assumptions, censoring at random, may have been violated due to the high level of non-CV event mortality competing risk (73.4 events per 1000 person-years follow-up) compared to CV events (31.1 per 1000 person-years). Such an example of how this issue has been addressed is through the CKD Prognosis Consortium’s “low eGFR events” model (22). We were unable to externally validate this model as it also included endstage kidney disease as an outcome and the correct method for model validation is currently unclear.

In summary the current manuscript describes an external CKD specific and general population CV risk prediction models in a large CKD cohort with a high number of robustly identified CV outcomes. CKD specific models generally performed poorly and general population models overpredicted risk, likely due to a high level of non-CV mortality competing risk events. Implementation of CV risk prediction models into clinical decision making in CKD should be cautiously considered and include the competing risk of other events such as all-cause mortality. Therefore, there is currently no reason to change the conclusion of Tangri *et* al’s previous systematic review that ‘further development of models for cardiovascular events [in CKD]…….is needed’ (3).

## Data Availability

All data produced in the present study are available upon reasonable request to the authors.

## Acknowledgements and Funding Statement

The authors would like to thank Dr Navdeep Tangri for providing literature review search strategy from the original systematic review. The author would also like to thank Mr Keith Nockels, Clinical Librarian, University of Leicester, for assisting with reviewing the literature review strategy.

RM was funded by Kidney Research UK (Grant TF2/2015) to develop the LCC cohort and investigate the topic of the manuscript. Other than external peer review of the grant for the study, Kidney Research UK had no role in study design; collection, analysis, and interpretation of data; writing the report; or the decision to submit the current report for publication.

