## Supplementary for "Cardiovascular Risk Models in Chronic Kidney Disease: A Systematic Review and External Validation of Models"

**Supplementary Item 1:** PRISMA checklist for the manuscript.

**Supplementary Item 2:** TRIPOD checklist for the manuscript.

**Supplementary Item 3:** Literature Search Strategy.

**Supplementary Figure 1**: Literature Review and Assessment of CKD Risk Models.

**Supplementary Table 1:** KDIGO CKD stages based on categorisation of EPI eGFR and ACR.

**Supplementary Table 2:** Summary C-statistics for CKD and General Population CV prediction models for complete case analysis.

**Supplementary Table 3:** Sensitivity Analysis for C-Statistics for QRisk3 Models**.**

**Supplementary Item 1: PRISMA 2009 Checklist**

| **Section/topic** | **#** | **Checklist item** | **Reported on page #** |
| --- | --- | --- | --- |
| **TITLE** | | |  |
| Title | 1 | Identify the report as a systematic review, meta-analysis, or both. | 1 |
| **ABSTRACT** | | |  |
| Structured summary | 2 | Provide a structured summary including, as applicable: background; objectives; data sources; study eligibility criteria, participants, and interventions; study appraisal and synthesis methods; results; limitations; conclusions and implications of key findings; systematic review registration number. | 2,3 |
| **INTRODUCTION** | | |  |
| Rationale | 3 | Describe the rationale for the review in the context of what is already known. | 4,5 |
| Objectives | 4 | Provide an explicit statement of questions being addressed with reference to participants, interventions, comparisons, outcomes, and study design (PICOS). | 5 |
| **METHODS** | | |  |
| Protocol and registration | 5 | Indicate if a review protocol exists, if and where it can be accessed (e.g., Web address), and, if available, provide registration information including registration number. | 6 & suppl |
| Eligibility criteria | 6 | Specify study characteristics (e.g., PICOS, length of follow-up) and report characteristics (e.g., years considered, language, publication status) used as criteria for eligibility, giving rationale. | 6 & suppl |
| Information sources | 7 | Describe all information sources (e.g., databases with dates of coverage, contact with study authors to identify additional studies) in the search and date last searched. | 6 & suppl |
| Search | 8 | Present full electronic search strategy for at least one database, including any limits used, such that it could be repeated. | 6 & suppl |
| Study selection | 9 | State the process for selecting studies (i.e., screening, eligibility, included in systematic review, and, if applicable, included in the meta-analysis). | 6 & suppl |
| Data collection process | 10 | Describe method of data extraction from reports (e.g., piloted forms, independently, in duplicate) and any processes for obtaining and confirming data from investigators. | 6 & suppl |
| Data items | 11 | List and define all variables for which data were sought (e.g., PICOS, funding sources) and any assumptions and simplifications made. | 6 & suppl |
| Risk of bias in individual studies | 12 | Describe methods used for assessing risk of bias of individual studies (including specification of whether this was done at the study or outcome level), and how this information is to be used in any data synthesis. | N/A |
| Summary measures | 13 | State the principal summary measures (e.g., risk ratio, difference in means). | 7,8 |
| Synthesis of results | 14 | Describe the methods of handling data and combining results of studies, if done, including measures of consistency (e.g., I^2^) for each meta-analysis. | N/A |

| **Section/topic** | **#** | **Checklist item** | **Reported on page #** |
| --- | --- | --- | --- |
| Risk of bias across studies | 15 | Specify any assessment of risk of bias that may affect the cumulative evidence (e.g., publication bias, selective reporting within studies). | N/A |
| Additional analyses | 16 | Describe methods of additional analyses (e.g., sensitivity or subgroup analyses, meta-regression), if done, indicating which were pre-specified. | N/A |
| **RESULTS** | | |  |
| Study selection | 17 | Give numbers of studies screened, assessed for eligibility, and included in the review, with reasons for exclusions at each stage, ideally with a flow diagram. | 9, Table 1 |
| Study characteristics | 18 | For each study, present characteristics for which data were extracted (e.g., study size, PICOS, follow-up period) and provide the citations. | 10, Table 2 |
| Risk of bias within studies | 19 | Present data on risk of bias of each study and, if available, any outcome level assessment (see item 12). | N/A |
| Results of individual studies | 20 | For all outcomes considered (benefits or harms), present, for each study: (a) simple summary data for each intervention group (b) effect estimates and confidence intervals, ideally with a forest plot. | N/A |
| Synthesis of results | 21 | Present results of each meta-analysis done, including confidence intervals and measures of consistency. | N/A |
| Risk of bias across studies | 22 | Present results of any assessment of risk of bias across studies (see Item 15). | N/A |
| Additional analysis | 23 | Give results of additional analyses, if done (e.g., sensitivity or subgroup analyses, meta-regression [see Item 16]). | suppl |
| **DISCUSSION** | | |  |
| Summary of evidence | 24 | Summarize the main findings including the strength of evidence for each main outcome; consider their relevance to key groups (e.g., healthcare providers, users, and policy makers). | 14,15 |
| Limitations | 25 | Discuss limitations at study and outcome level (e.g., risk of bias), and at review-level (e.g., incomplete retrieval of identified research, reporting bias). | 15,16 |
| Conclusions | 26 | Provide a general interpretation of the results in the context of other evidence, and implications for future research. | 16,17 |
| **FUNDING** | | |  |
| Funding | 27 | Describe sources of funding for the systematic review and other support (e.g., supply of data); role of funders for the systematic review. | 18 |

**Supplementary Item 2 - TRIPOD Checklist**

| **Section/Topic** | **Item** | **Checklist Item** | **Page** |
| --- | --- | --- | --- |
| **Title and abstract** | | | |
| Title | 1 | Identify the study as developing and/or validating a multivariable prediction model, the target population, and the outcome to be predicted. | 1 |
| Abstract | 2 | Provide a summary of objectives, study design, setting, participants, sample size, predictors, outcome, statistical analysis, results, and conclusions. | 2,3 |
| **Introduction** | | | |
| Background and objectives | 3a | Explain the medical context (including whether diagnostic or prognostic) and rationale for developing or validating the multivariable prediction model, including references to existing models. | 4,5 |
|  | 3b | Specify the objectives, including whether the study describes the development or validation of the model or both. | 5 |
| **Methods** | | | |
| Source of data | 4a | Describe the study design or source of data (e.g., randomized trial, cohort, or registry data), separately for the development and validation data sets, if applicable. | 6 |
|  | 4b | Specify the key study dates, including start of accrual; end of accrual; and, if applicable, end of follow-up. | 6 |
| Participants | 5a | Specify key elements of the study setting (e.g., primary care, secondary care, general population) including number and location of centres. | 6 |
|  | 5b | Describe eligibility criteria for participants. | 6 |
|  | 5c | Give details of treatments received, if relevant. | N/A |
| Outcome | 6a | Clearly define the outcome that is predicted by the prediction model, including how and when assessed. | 6 |
|  | 6b | Report any actions to blind assessment of the outcome to be predicted. | N/A |
| Predictors | 7a | Clearly define all predictors used in developing or validating the multivariable prediction model, including how and when they were measured. | 7 |
|  | 7b | Report any actions to blind assessment of predictors for the outcome and other predictors. | N/A |
| Sample size | 8 | Explain how the study size was arrived at. | N/A |
| Missing data | 9 | Describe how missing data were handled (e.g., complete-case analysis, single imputation, multiple imputation) with details of any imputation method. | 7 |
| Statistical analysis methods | 10c | For validation, describe how the predictions were calculated. | 8 |
|  | 10d | Specify all measures used to assess model performance and, if relevant, to compare multiple models. | 8 |
|  | 10e | Describe any model updating (e.g., recalibration) arising from the validation, if done. | 8 |
| Risk groups | 11 | Provide details on how risk groups were created, if done. | 8 |
| Development vs. validation | 12 | For validation, identify any differences from the development data in setting, eligibility criteria, outcome, and predictors. | 7,8  Table 2 |
| **Results** | | | |
| Participants | 13a | Describe the flow of participants through the study, including the number of participants with and without the outcome and, if applicable, a summary of the follow-up time. A diagram may be helpful. | 11  Table 3 |
|  | 13b | Describe the characteristics of the participants (basic demographics, clinical features, available predictors), including the number of participants with missing data for predictors and outcome. | 11  Table 3 |
|  | 13c | For validation, show a comparison with the development data of the distribution of important variables (demographics, predictors and outcome). | 11 |
| Model performance | 16 | Report performance measures (with CIs) for the prediction model. | 12,13  Table 4, Figures |
| Model-updating | 17 | If done, report the results from any model updating (i.e., model specification, model performance). | 13, Figures |
| **Discussion** | | | |
| Limitations | 18 | Discuss any limitations of the study (such as nonrepresentative sample, few events per predictor, missing data). | 15,16 |
| Interpretation | 19a | For validation, discuss the results with reference to performance in the development data, and any other validation data. | 14,15,16 |
|  | 19b | Give an overall interpretation of the results, considering objectives, limitations, results from similar studies, and other relevant evidence. | 14,15,16 |
| Implications | 20 | Discuss the potential clinical use of the model and implications for future research. | 16,17 |
| **Other information** | | | |
| Supplementary information | 21 | Provide information about the availability of supplementary resources, such as study protocol, Web calculator, and data sets. | Suppl |
| Funding | 22 | Give the source of funding and the role of the funders for the present study. | 18 |

**Supplementary Item 3 – Literature Search Strategy**

1 chronic kidney disease.tw.

2 chronic kidney disorder.tw.

3 kidney failure, chronic/ or chronic kidney failure.tw.

4 chronic kidney insufficiency.tw.

5 chronic nephropathy.tw.

6 chronic renal disease.tw.

7 chronic renal failure.tw.

8 renal insufficiency, chronic/ or chronic renal insufficien$.tw.

9 1 or 2 or 3 or 4 or 5 or 6 or 7 or 8

10 predict*.tw.

11 validat*.tw.

12 develop*.tw.

13 Predictive value of tests/

14 scor*.tw.

15 observ*.tw.

16 Observer variation/

17 ROC curve/

18 discriminat*.tw.

19 c-statistic.tw.

20 c statistic.tw.

21 area under the curve.tw.

22 area under curve.tw.

23 auc.tw.

24 calibration.tw.

25 Algorithm/

26 10 or 11 or 12 or 13 or 14 or 15 or 16 or 17 or 18 or 19 or 20 or 21 or 22 or 23

or 24 or 25

27 Mortality/

28 (mortal$ or dead or death).tw.

29 cardiovascular disease.tw. or Cardiovascular Diseases/

30 exp Cardiovascular Diseases/

31 cardi*.tw.

32 heart.tw.

33 myocard*.tw.

34 ischem*.tw.

35 ischaem*.tw.

36 stroke.tw.

37 cerebrovasc*.tw.

38 cerebral vascular*.tw.

39 endstage renal.tw.

40 endstage kidney.tw.

41 esrf.tw.

42 esrd.tw.

43 transplant*.tw.

44 peritoneal dialysis.tw.

45 hemodialysis.tw.

46 haemodialysis.tw.

47 27 or 28 or 29 or 30 or 31 or 32 or 33 or 34 or 35 or 36 or 37 or 38 or 39 or 40

or 41 or 42 or 43 or 44 or 45 or 46

48 9 and 26 and 47

49 limit 48 to (english language and humans and yr="2012 -Current")

50 limit 49 to (clinical study or clinical trial, all or clinical trial or comparative study

or evaluation studies or observational study or validation studies)

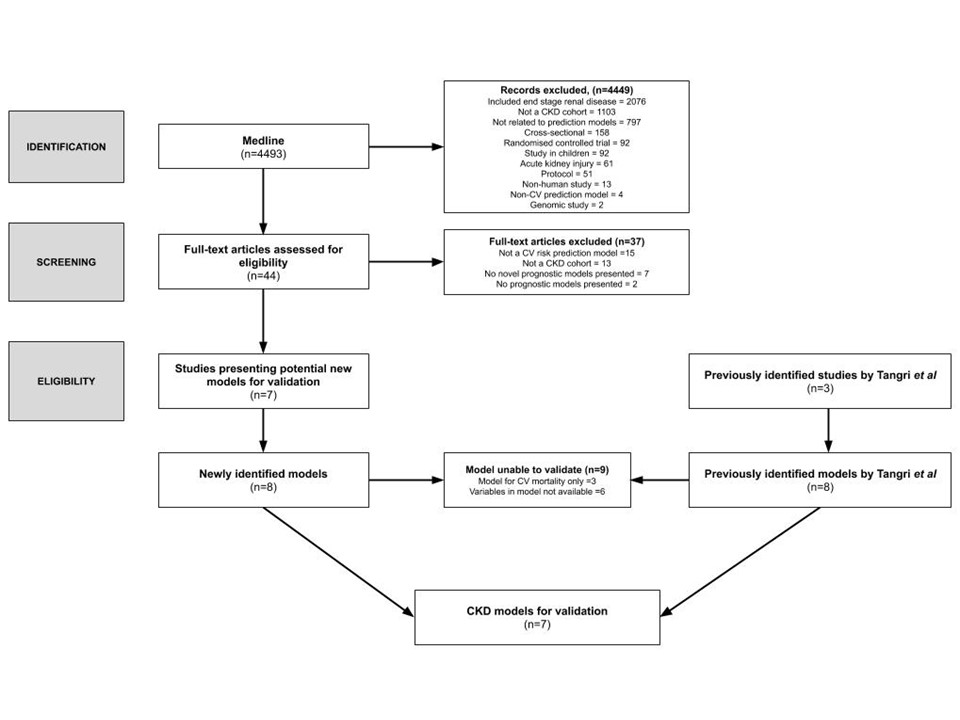

**Supplementary Figure 1**: Literature Review and Assessment of CKD Risk Models.

|  | **ACR Stage** | | | |  |
| --- | --- | --- | --- | --- | --- |
| **EPI Stage** | **Missing** | **1**  **(ACR<3)** | **2**  **(ACR 3-30)** | **3**  **ACR >30)** | **Total** |
| **3A (eGFR 45-59)** | **4,657** | **4,266** | **3,169** | **270** | **12,362** |
| % | 27.0 | 24.7 | 18.4 | 1.6 | 71.7 |
| **3B (eGFR 30-44)** | **801** | **1,386** | **1,486** | **250** | **3,923** |
| % | 4.6 | 8.0 | 8.6 | 1.5 | 22.7 |
| **4 (eGFR 15-29)** | **140** | **189** | **367** | **173** | **869** |
| % | 0.8 | 1.1 | 2.1 | 1.0 | 5.0 |
| **5 (eGFR <15)** | **17** | **5** | **27** | **45** | **94** |
| % | 0.1 | 0.03 | 0.2 | 0.3 | 0.5 |
| **Total** | **5,615** | **5,846** | **5,049** | **738** | **17,248** |
| % | 32.6 | 33.9 | 29.3 | 4.3 |  |

**Supplementary Table 1**: KDIGO CKD stages based on categorisation of EPI eGFR and ACR. All eGFR units in ml/min/1.73m^2^ and ACR units in mg/mmol

| **Model** | **Original Performance** | | **Model Specific** | | **Whole Cohort** | |
| --- | --- | --- | --- | --- | --- | --- |
|  | **C** | **95% CI** | **C** | **95% CI** | **C** | **95% CI** |
| **CKD** |  |  |  |  |  |  |
| Alderson | 0.873 | 0.825 to 0.921 | n/a | n/a | 0.587 | 0.575 to 0.600 |
| Weiner Male Framingham | 0.62 | not reported | 0.606 | 0.570 to 0.641 | 0.586 | 0.567 to 0.605 |
| Weiner Male Best Cox | 0.72 | not reported | 0.557 | 0.520 to 0.593 | 0.560 | 0.540 to 0.579 |
| Weiner Female Framingham | 0.77 | not reported | 0.567 | 0.535 to 0.598 | 0.563 | 0.545 to 0.580 |
| Weiner Female Best Cox | 0.82 | not reported | 0.549 | 0.517 to 0.582 | 0.540 | 0.522 to 0.558 |
| Matsushita Male Framingham | 0.679 | not reported | 0.640 | 0.606 to 0.674 | 0.588 | 0.569 to 0.607 |
| Matsushita Female Framingham | 0.679 | not reported | 0.611 | 0.586 to 0.636 | 0.570 | 0.581 to 0.614 |
| **General Population** |  |  |  |  |  |  |
| QRisk2 Male | 0.792 | 0.789 to 0.794 | 0.637 | 0.574 to 0.699 | 0.625 | 0.606 to 0.644 |
| QRisk2 Female | 0.817 | 0.814 to 0.820 | 0.680 | 0.639 to 0.720 | 0.676 | 0.660 to 0.691 |
| QRisk3 Male | 0.858 | 0.857 to 0.860 | 0.640 | 0.542 to 0.739 | 0.608 | 0.582 to 0.634 |
| QRisk3 Female | 0.880 | 0.879 to 0.882 | 0.695 | 0.643 to 0.747 | 0.664 | 0.644 to 0.683 |
| PCE Male | 0.746 | not reported | 0.663 | 0.611 to 0.714 | 0.616 | 0.597 to 0.635 |
| PCE Female | 0.806 | not reported | 0.645 | 0.607 to 0.684 | 0.665 | 0.650 to 0.680 |

**Supplementary Table 2: Summary C-statistics for CKD and General Population CV prediction models.** C-statistics and 95% CI presented are for complete case analysis. C – Harrell’s concordance statistic, CI – confidence interval.

|  | **Female** | | | | **Male** | | | |
| --- | --- | --- | --- | --- | --- | --- | --- | --- |
|  | **Model Specific** | | **Whole Cohort** | | **Model Specific** | | **Whole Cohort** | |
|  | **C** | **95% CI** | **C** | **95% CI** | **C** | **95% CI** | **C** | **95% CI** |
| All CVD events | 0.684 | 0.647 to 0.720 | 0.666 | 0.651 to 0.681 | 0.628 | 0.573 to 0.683 | 0.618 | 0.599 to 0.637 |
| **Age Group** |  |  |  |  |  |  |  |  |
| <59 | 0.792 | 0.586 to 0.998 | 0.712 | 0.583 to 0.840 | 0.571 | 0.269 to 0.873 | 0.591 | 0.476 to 0.705 |
| 60-69 | 0.578 | 0.373 to 0.783 | 0.696 | 0.625 to 0.767 | 0.455 | 0.306 to 0.605 | 0.591 | 0.529 to 0.652 |
| 70-79 | 0.559 | 0.483 to 0.635 | 0.581 | 0.550 to 0.613 | 0.589 | 0.495 to 0.684 | 0.602 | 0.571 to 0.634 |
| 80+ | 0.566 | 0.505 to 0.627 | 0.580 | 0.560 to 0.601 | 0.568 | 0.486 to 0.651 | 0.554 | 0.526 to 0.582 |
| **Outcomes** |  |  |  |  |  |  |  |  |
| MI Only | 0.668 | 0.603 to 0.734 | 0.666 | 0.643 to 0.690 | 0.562 | 0.462 to 0.661 | 0.611 | 0.584 to 0.638 |
| Stroke Only | 0.688 | 0.646 to 0.730 | 0.668 | 0.650 to 0.685 | 0.672 | 0.604 to 0.741 | 0.621 | 0.597 to 0.646 |
| Death Only | 0.675 | 0.651 to 0.699 | 0.701 | 0.691 to 0.710 | 0.651 | 0.617 to 0.685 | 0.655 | 0.643 to 0.666 |
| MI, Stroke & Death | 0.677 | 0.656 to 0.698 | 0.687 | 0.678 to 0.696 | 0.642 | 0.611 to 0.674 | 0.640 | 0.629 to 0.651 |
| Addition of eGFR and ACR* | 0.010 | -0.026 to 0.047 | 0.018 | 0.003 to 0.033 | 0.021 | -0.033 to 0.076 | 0.016 | -0.003 to 0.034 |
| Complete Cases Only | 0.646 | 0.629 to 0.664 | 0.664 | 0.648 to 0.679 | 0.608 | 0.588 to 0.629 | 0.617 | 0.598 to 0.636 |

**Supplementary Table 3: Sensitivity Analysis for C-Statistics for QRisk3 Models.** C and 95% CI presented are the mean of C results for each imputed cycle. *change in C based on recalculated risk of models with the addition of log transformed eGFR and ACR as additional continuous predictor variables. C – Harrell’s concordance statistic, CI – confidence interval.
